# Impact of direct-to-consumer genetic testing on Australian clinical genetics services

**DOI:** 10.1101/2020.03.05.20031963

**Authors:** Michael Millward, Jane Tiller, Michael Bogwitz, Helen Kincaid, Shelby Taylor, Alison H Trainer, Paul Lacaze

## Abstract

**Purpose:** The increasing popularity of direct-to-consumer genetic testing (DTCGT) is thought to be creating a burden on clinical genetic health services worldwide. However, no studies have collected recent evidence regarding the extent of this impact in Australia.

**Methods:** We administered an online survey to Australian clinical genetics services, asking questions related to DTCGT-related referrals received and outcomes over the past 10 years.

**Results:** Eleven publicly-funded clinical genetics services completed the survey, reporting over 100 DTCGT-related referrals. Most referrals (83%) were made by general practitioners seeking interpretation of DTCGT results. More than 30% of referrals related to imputed genetic risk estimates generated from third-party web-based software tools. Services reported low validation rates for DTCGT results (<10%). Procedures for managing DTCGT referrals and granting appointments were variable between services, with most services (8/11) lacking specific procedures.

**Conclusion:** Our study helps quantify the impact of DTCGT on clinical genetics services, and highlights the impact of imputed genetic risk estimates generated from third-party software.

## Introduction

Direct-to-consumer genetic tests (DTCGT) are DNA tests sold directly to consumers, without the involvement of a medical professional ^1,2^. DTCGTs can include health-related DNA testing, or testing related to ancestry or other traits. Many DTCGTs are not required to adhere to the same regulations as accredited medical genetic testing in given jurisdictions ^1^, sometimes resulting in inaccurate or misleading reporting of results ^3,4^.

Another popular aspect of DTCGT is consumers retrieving raw genotype data from DTCGT providers, then uploading it into third-party web-based interpretation software tools ^5^ to generate imputed genetic risk estimates for diseases ^6^. Unfortunately, these risk estimates are often not accurate, and have a high propensity for false positive results ^6^. A recent study attempted to validate DTCGT results in a clinically-accredited lab, and found that 40% of variants reported across a selection of patient samples were false positives ^3^. Additionally, the health reports and risk estimates produced by these third-party software imputation tools can be difficult for many consumers and healthcare professionals to interpret, which at times can create concern ^6^.

Given the growing popularity of DTCGT and issues related of technical validity and interpretation ^4,7^, clinical genetics services are likely to receive more DTCGT-related referrals. Concerns have arisen internationally regarding the likely public healthcare costs that will flow from consumers seeking follow-up for DTCGT and information derived from third-party imputation tools^8^. However, data on the current clinical impact of this is lacking^9^.

In Australia and other countries, limited state funding supports genetic health services and is used to prioritize patient appointments and the use of clinical genetic tests with proven clinical utility^10^. Many services are under-resourced, resulting in long waiting times for appointments, and other challenges ^11^. We hypothesise that the growing popularity of DTCGT within Australia is resulting in more referrals, creating a further burden. However, no recently published research has been undertaken to confirm whether this is the case, and if so, the extent of the burden. Here, we collect some of the first evidence to quantify the impact of DTCGT-related referrals on clinical genetics services in Australia.

## Materials and methods

An online REDCap survey was co-designed with experienced genetic health professionals in Australia, to collect information on DTCGT-related referrals received over the last 10 years. The survey questions were separated into four sections which collected data regarding the genetics service; i) the estimated number of referrals received, ii) clinical actions typically following DTCGT- related referrals; iii) the types of DTCGT-related referrals received; and iv) details of procedures for managing DTCGT-related referrals (for a complete list of questions included in the survey, see Supplementary Materials). Nineteen publicly-funded genetics services (those known to be currently operating in Australia) were invited to participate. The survey was anonymous, except that it asked for the Australian state in which each service is based. Descriptive data analysis was performed using STATA. Ethical approval for this project was received from The Royal Children’s Hospital Melbourne Human Research Ethics Committee.

## Results

Of the 19 services invited to participate, 13 survey responses were received. Two responses did not contain substantive data as they did not progress beyond the initial questions. They were excluded from analysis, leaving 11 eligible responses. Data were collected from at least one service in the majority of states and territories.

The number of DTCGT-related referrals varied between services (Figure 1A). Some services (n=3/9) estimated receiving only 1-5 DTCGT-related referrals in the past 10 years, while others (n=2/9) estimated receiving 31-35 in the same time period. Two services did not provide an estimate, but both indicated by free text response that they had received DTCGT-related referrals in the last 10 years. Based on these estimates, at least 114 DTCGT-related referrals have been received by Australian genetics services since 2010.

**Figure 1:**
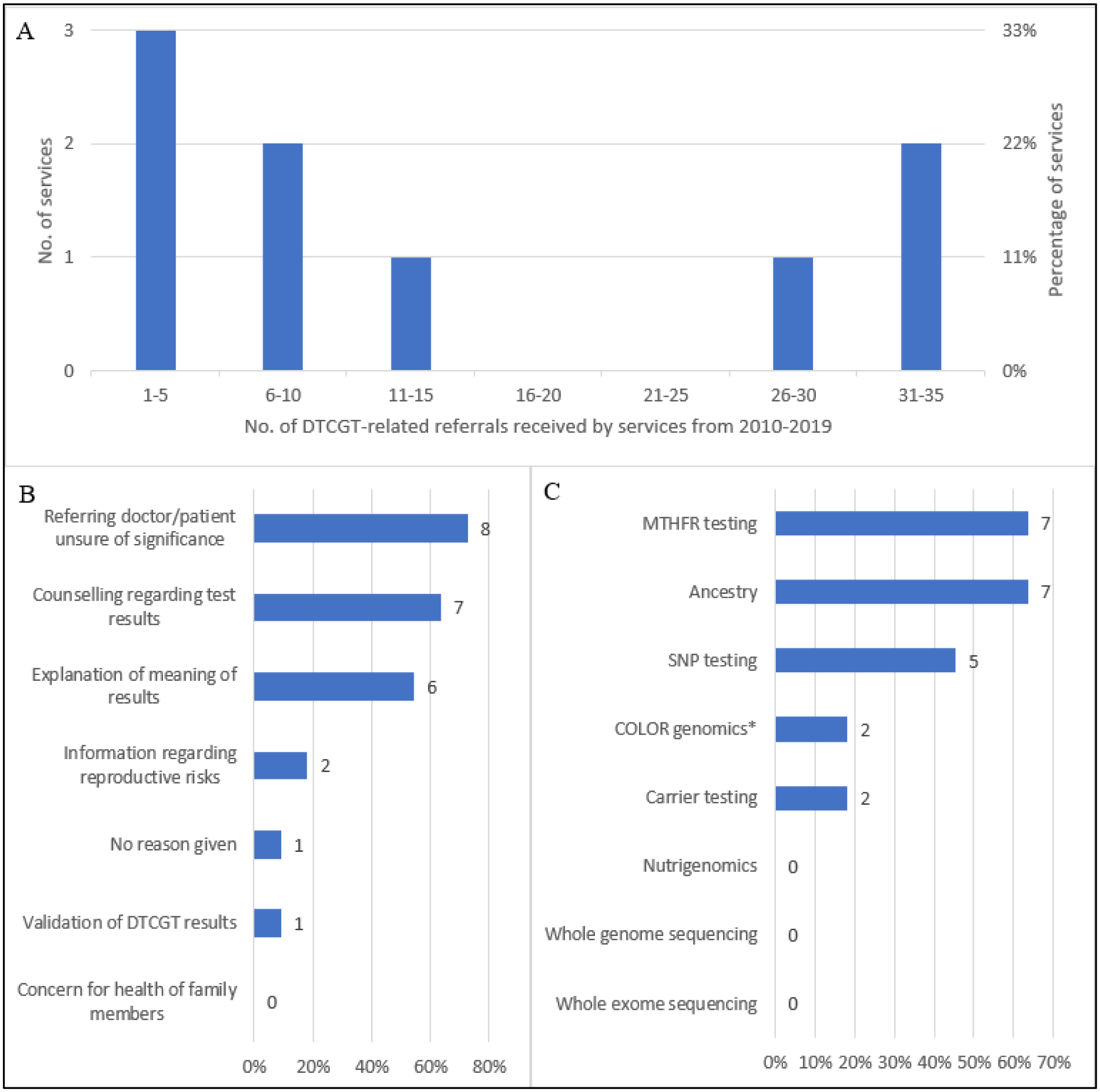
A) Estimated no. of DTCGT-related referrals received by public genetics services in Australia from 2010-2019 (n=9). B) Referral reasons most commonly associated with DTCGT-related referrals (n=11). The value at the end of each bar represents the total number of PGS that selected this option. PGS could select more than one option. C) DTCGT type most commonly associated with DTCGT-related referrals (n=11). The value at the end of each bar represents the total number of PGS that selected this option. Abbreviations: DTCGT = direct-to-consumer genetic testing.

When asked about the ‘reasons’ for receiving DTCGT-related referrals, seven different reasons were reported (Figure 1B). The most common reason was ‘the referrer being unsure of the significance of the DTCGT results’ (n=8/11). *Methylenetetrahydrofolate reductase (MTHFR)* gene testing and ancestry testing were the equal most common types of testing associated with DTCGT-related referrals (n=7/11, Figure 1C).

Many services (50%) reported receiving referrals for consumers who had generated imputed risk estimates from third-party web-based software tools. Services estimated that, on average, 33% of DTCGT-related referrals related to results obtained using these third-party tools.

When asked to estimate on a sliding scale, services reported over 50% (52.2%) of DTCGT-related referrals resulted in patient appointments. However, the willingness to offer appointments varied between services. Three services gave zero appointments to patients for DTCGT-related referrals, and three gave appointments for all DTCGT-related referrals. The remaining three services saw a proportion of patients. Following an appointment for a DTCGT-related referral, Figure 2A shows the number of services that reported specific additional clinical actions (green) and the number of individual patients across all services that required those specific clinical actions (orange).

**Figure 2:**
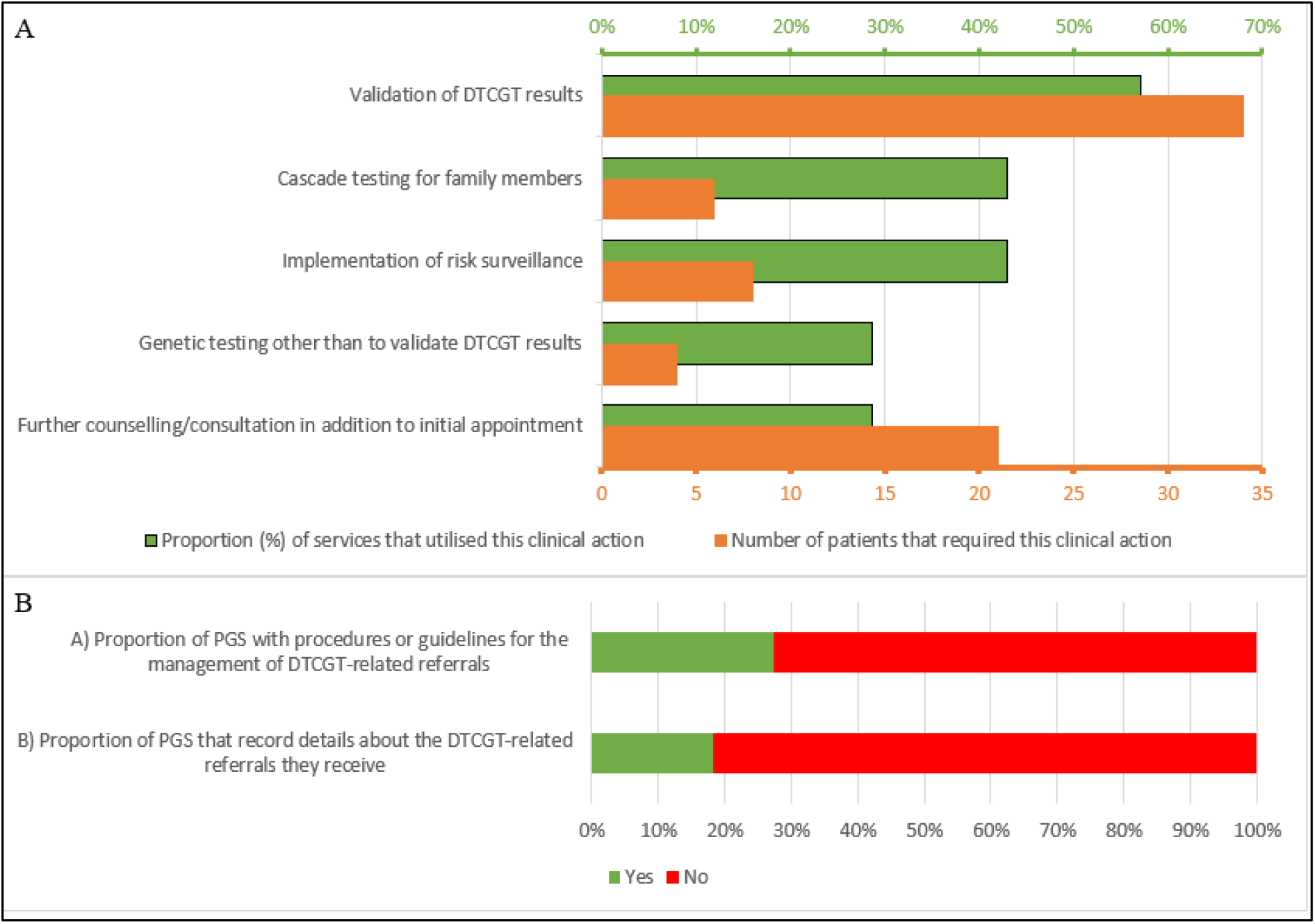
A) Additional clinical actions required following patient appointments resulting from DTCGT-related referrals (n=11). B) Procedures for the management and collection of information regarding DTCGT-related referrals (n=11). Abbreviations: DTCGT = direct-to-consumer genetic testing.

Over 50% of services (n=6/11) reported the validation of DTCGT results to be the most common clinical action following an appointment (Figure 2A). In total, the services surveyed estimated attempting to validate 34 patients’ results. However, only 3 (9%) of these results were shown to be correct.

Most services (n=8/11) had no specific procedures in place for managing or recording details of DTCGT-related referrals they received (n=9/11) (Figure 2B). Two services specified procedures for managing referrals associated with *MTHFR* gene variants specifically.

## Discussion

Our study undertook a survey of clinical genetics services across Australia, to determine the impact of DTCGT-related referrals on the delivery of health-related services. The results generated by the study provides some of the first evidence of a tangible impact of DTCGT on health services related to genetics.

Our study produced a number of notable results. First, Australian genetics services reported low rates of validation for DTCGT results (<10%). This validation rate was even lower than previously reported studies ^3^. Publicly-funded genetics services have limited testing budgets in Australia; meaning decisions to pay for validation testing of DTCGT are necessarily restricted by the limited capacity to pay for other types of genetic testing that may be deemed more necessary. A high proportion of false-positive DTCGT results represents a resource burden on these services. It also speaks to the potential for negative or unnecessary concern generated for consumers, and time wasted, as a result of false-positive results^9^.

Second, we found that Australian genetics services reported more than 30% of all DTCGT-related referrals related to imputed risk estimates generated by consumers using third-party web-based software tools. To our knowledge, this provides some of the first evidence of the impact caused by third-party tools, compared with the impact of original DTCGT results on clinical genetics services. It also highlights the unintended consequences that can flow from DTCGT providers’ return of raw genotype data to consumers. In most cases, consumers are entitled to request their raw data, despite the subsequent risks. Our findings suggest more stringent regulation or validation protocols may be required with regards to the use of third-party genetic risk-imputation software tools^12^, alongside better public education to help promote awareness of the limitations of these tools.

Third, we collected data on post-appointment clinical actions, including risk-surveillance and cascade testing of family members, after validation of DTCGT results. Almost half of Australian services reported taking clinical actions for at least one DTCGT-related referral. This demonstrates that when DTCGT-related results are technically valid, appropriate downstream clinical management can follow. However, the proportion of validated DTCGT results, compared with false-positives, was very low.

Fourth, regarding reasons for genetics services receiving DTCGT-related referrals, we found that the majority (84%, n=21/25) of reasons related to DTCG result misinterpretation, either by the consumer and/or the referring doctor. This is consistent with previous studies ^13,14^, and suggests consumers and referring doctors still lack confidence in their ability to correctly interpret DTCGT results ^15,16^.

Fifth, we found that the most common types of DTCGT-referrals reported related to *MTHFR* variants (n=7/11) and SNP profiles (n=5/11) – both commonly associated with mis-interpretation ^17,18^. For example, some DTCGT services associate *MTHFR* gene variants with a range of conditions including autism, cardiac disease, and poor pregnancy outcomes, for which there is limited supporting evidence ^17^.

Sixth, we found considerable variation in the way different clinical genetics services manage DTCGT- related referrals, with no common procedure or policy in place. This may reflect variation in the services’ perception of DTCGT utility ^14^, demand from other areas, and/or differences in resources. We suggest that a consistent approach, supported by policy or guidelines for managing DTCGT- related referrals, is required.

### Limitations

Study limitations include not all of the Australian public genetics services responding to our survey, meaning the number of DTCGT-related referrals nation-wide is likely underestimated. The survey was anonymous, meaning services could not be followed-up to ask more specific questions, or compare the size of each responding service with number of relative referrals received. This limited our ability to determine geographic areas with higher numbers of DTCGT-related referrals. As services were asked to estimate responses, accuracy of estimates may have been limited by the service representatives’ knowledge and/or perspective on DTCGT.

Our study provides important new evidence regarding the impact of DTCGT on genetics services in Australia. From here, we suggest a more comprehensive and formal audit is required, which can be repeated periodically, to further quantify and understand the impact of DTCGT across all Australian genetics services. Our study also provides a platform for further international studies exploring the burden of DTCGT and data obtained from third-party imputation software on local genetics services.

## Conclusion

Our study has confirmed, as suspected, that the increasing popularity of DTCGT is causing a material impact on publicly-funded Australian genetics services. We also report a sizeable portion of that impact is resulting from the use of third-party genetic risk-imputation software tools. The impact of DTCGT is likely to increase in coming years, if the use of DTCGT continues to grow.

## Data Availability

Supplementary data (copy of online survey) is uploaded for review

## Acknowledgements

This study was completed in partial fulfilment of the requirements for the Master of Genetic Counselling, University of Melbourne, Victoria, Australia, and was supported by the Victorian Government’s Operational Infrastructure Support Program.

